# Early Warning of Atrial Fibrillation Using Deep Learning

**DOI:** 10.1101/2022.09.05.22279605

**Authors:** Marino Gavidia, Hongling Zhu, Arthur N. Montanari, Jesús Fuentes, Cheng Cheng, Sergio Dubner, Martin Chames, Pierre Maison-Blanche, Md Moklesur Rahman, Roberto Sassi, Fabio Badilini, Yinuo Jiang, Shengjun Zhang, Hai-Tao Zhang, Hao Du, Basi Teng, Ye Yuan, Guohua Wan, Zhouping Tang, Xin He, Xiaoyun Yang, Jorge Goncalves

**Author notes:** Equal contributor (Marino Gavidia, Hongling Zhu).

## Abstract

Atrial Fibrillation (AF) is the most common cardiac rhythm disorder. It is associated with increased hospitalization, risk of heart failure, and risk of stroke. Once on AF, it can sometimes be difficult to revert to sinus rhythm (SR), potentially requiring pharmacological or electrical cardioversion. Earlier warning of an imminent switch from SR to AF, even if by only a few minutes, could prompt patients to take actions (e.g., taking oral antiarrhythmic drugs) to avoid AF and its associated complications, thereby easing the workload and costs to the health system. The question is whether there is information, even if subtle, in the minutes prior to AF to indicate an imminent switch from SR. This paper shows that, for the vast majority of patients, the answer is affirmative. We propose a deep-learning model that predicts the onset of AF on average 30.8 minutes before it appears, with an accuracy of 83% and an F1-score of 85% on the test data. Moreover, this performance was obtained from R-to-R interval signals, which can be obtained from common wearable devices such as smartwatches and smart bands. The predictions were performed using a deep convolutional neural network, trained and cross-validated on 24-hour data obtained from Holter electrocardiogram recordings of 280 patients, with an additional 70 patients used as test data. We further tested the model with data from two other external centers with 33 patients. Overall, the proposed method has low computational time and could be embedded in smartphones connected to wearable devices that capture RRI for continuous heart monitoring and early warning of AF onset.

## Introduction

Atrial fibrillation (AF) is the most common cardiac arrhythmia worldwide; the estimated number of individuals with AF in 2010 was 33.5 million [1]. Hand in hand with the growing prevalence of AF [2], healthcare costs continue to increase mainly because of hospitalization and treatment costs [3]. AF episodes contribute to emergency department presentations due to high symptom burden and heart failure decompensation from tachycardiomyopathy. Maintaining sinus rhythm (SR) is a priority since AF events can increase the risk of other diseases [4], such as stroke and dementia [5, 6], as well as lead to atrial remodeling, which may enhance susceptibility to future episodes [7]. The early prediction of AF episodes in patients with paroxysmal AF could prompt patients to take preventive measures to maintain SR (e.g., avoid alcohol consumption or take preventive antirrhythmic and anticoagulation medication), possibly reducing emergency department presentations and associated healthcare costs. However, the identification of patients with a high likelihood of AF onset and its early-warning prediction (in the timescale of minutes or hours) are challenging problems in the clinical setting [8, 9]. To overcome these challenges, we develop a deep-learning model that continuously monitor patients to provide early warnings of imminent AF onsets. Such model could warn patients to take preventive medications only when really needed, instead of taking them regularly.

The automated *detection* of AF regimes from recorded electrocardiagram (ECG) data is a well-studied problem in the literature [10, 11]. Recent approaches based on machine learning and neural networks have achieved over 99% accuracy in the classification task [12–15], which led to functional applications on wearable devices of Apple, Fitbit, Samsung, and others [16–18]. On the other hand, the *prediction* of the onset of AF is still an open problem [19]. Numerous studies have developed models for long-term *risk assessments* of atrial fibrillation and other cardiovascular diseases, providing estimates typically on the order of weeks, months, or years [20–24]. Such machine-learning models for AF detection and risk assessment are often trained on short-duration ECG samples obtained from sporadic cardiologist controls. Although these datasets are very extensive (thousands or even millions of recordings), they do not typically contain long-duration ECG recordings (in the order of hours)—a type of data required for the development of models for real-time monitoring and prediction. Long-duration recordings require the inconvenient use of Holter devices or patches and are thus often collected from patients with more severe AF conditions. These factors substantially reduce the amount of data available for model training in forecasting problems for cardiovascular diseases.

Despite these data challenges, recent advances based on machine-learning and deep-learning models have been proposed for short-term prediction of AF using models trained on features extracted from ECG leads [25–28], R-to-R intervals (RRI) [29–33], or a combination of both [34–41]. All these methods have strong limitations. Most have limited data for model training—typically around 50 or less patients. More importantly, all methods use data up to, or very close to, the onset of AF to “predict” an AF event. Since little or no warning of AF onset is given in advance, these methods can be categorized as detection/classification tools rather than early warning models. As an example, Ref. [32] uses the entire window from four hours to zero hours (i.e., at onset) to classify whether an AF event will follow or not; in practice, it does not provide an early warning. In contrast, our work departs from this approach to a more prospective prediction model. Utilizing a sliding window feature, our model is designed to identify precursors of AF that are far away from the onset, thus providing early warnings on a timescale of minutes. Figure 1 illustrates the distinct data windows used by different models in detection, prediction at onset, and early-warning prediction.

**Figure 1:**
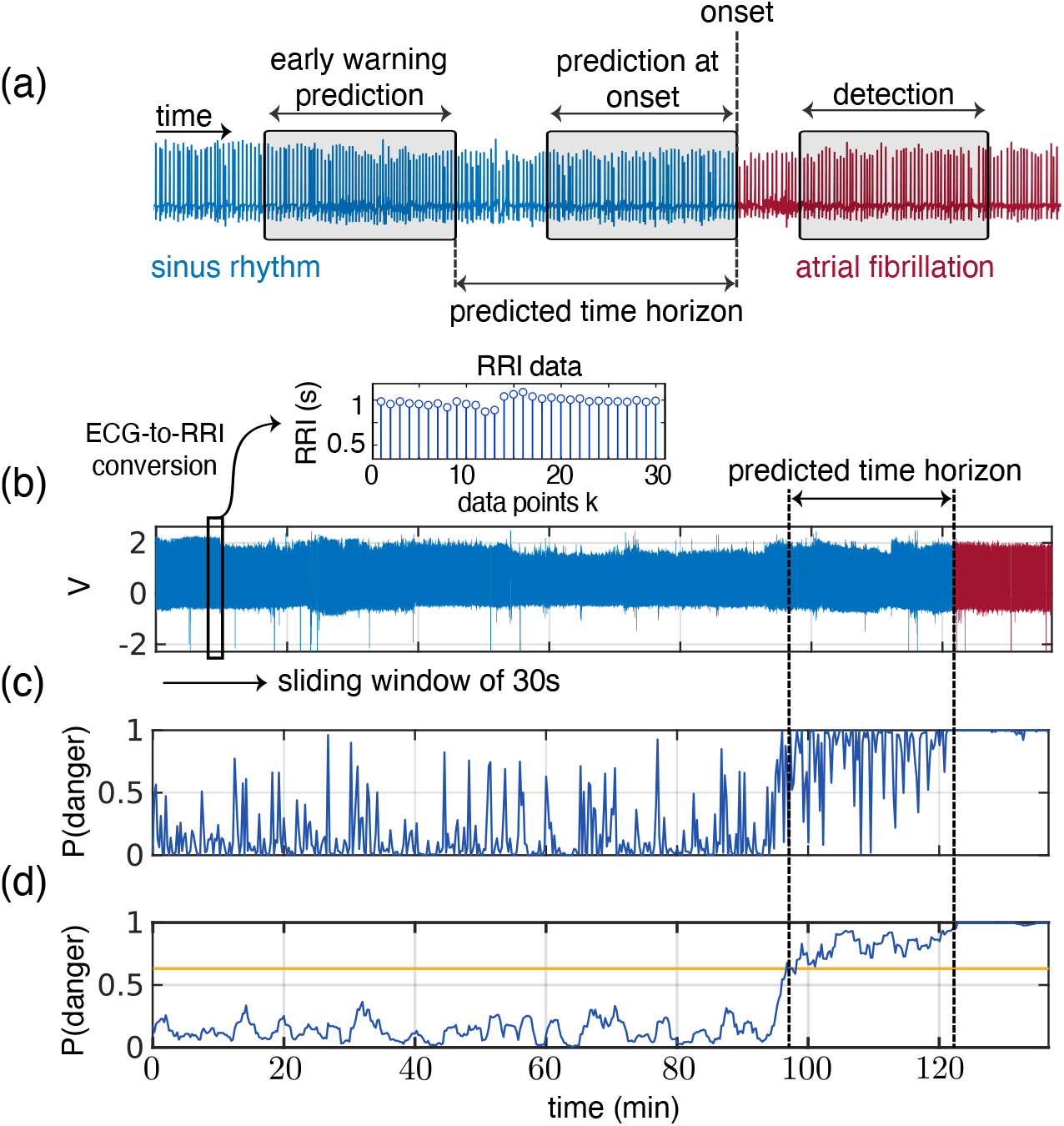
Detection versus prediction and early warnings. **(a)** Early-warning AF prediction (left box), AF prediction at onset (middle), and AF detection (right). The predicted time horizon is the time between the early warning and AF onset. **(b)** The model employs a sliding window, which sequentially samples the ECG data every 15 seconds. This window contains a duration of 30 seconds of ECG data, which is then converted to RRI data. Specifically, the total duration of the *n* RRI samples within the window is precisely 30 seconds. **(c)** For each sliding window of RRI data, the model computes the “probability of danger” to transition to AF. **(d)** Then, a non-anticipative moving average window smooths this probability of danger. When it crosses a threshold (yellow line), it triggers a warning of an imminent AF onset. For this particular patient, an alert is triggered 24min in advance of AF onset. Supplementary Movies illustrate the probability of danger rising as the AF onset approaches for other patients.

This paper presents a retrospective study that develops a deep-learning model for early warning of AF, entitled WARN (Warning of Atrial fibRilatioN). The model is trained and tested on 350 individual 24h-Holter recordings. On the test data, WARN gives early warnings of, on average, 30.8 minutes before onset of AF, with an accuracy and F1-score of 83% and 85%, respectively. Our model has a high performance using only RRI signals, which can be acquired from easy to wear and affordable pulse signal recorders, such as smartwatches or smart fitness bands. These devices can be used on a daily basis by patients, paving the way for real-time monitoring algorithms that learn and monitor long-term cardiac dynamics.

## Results

We used 24h-Holter recordings collected from 350 patients at Tongji Hospital (Wuhan, China) to develop and evaluate WARN for the early warning of AF. Recordings with short-duration AF episodes and/or significant noise artificats were excluded from the original dataset. The cohort was divided in two groups for the training/cross-validation (280 patients) and testing (70 patients) of WARN, as summarized in Table 1. The ECG recordings were converted to RRI data, as motivated by our ultimate goal of developing a monitoring/prediction method for wearable devices. Then, the RRI data were used to train a deep convolutional neural network (CNN) with 479 layers that classifies between SR, pre-AF, and AF segments. Finally, for each sliding window of 30s of RRI data, WARN outputs a “probability of danger” that the patient will have an imminent AF episode (Fig. 1b–d). Further dataset descriptions and method developments are described in Supplementary Material.

**Table 1:** Characteristic of the patients.

| Characteristic | Tongji Hospital, China |  | External centers |  |
| --- | --- | --- | --- | --- |
|  | Training cohort | Test cohort | France | Argentina |
| Total | 280 | 70 | 25 | 8 |
| Age < 65 | 115 (mean age 55) | 31 (mean age 54) | 8 (mean age 57) | 2 (mean age 60) |
| Age ≥ 65 | 165 (mean age 73) | 39 (mean age 73) | 17 (mean age 73) | 6 (mean age 80) |
| Male | 163 | 26 | 15 | 5 |
| Female | 117 | 44 | 10 | 3 |

We initially evaluated WARN on a test dataset consisting of RRI data. For comparison purposes, WARN was also trained and tested on ECG data, achieving slightly better performance. Finally, to evaluate WARN’s performance on out-of-sample data, we tested the model on 33 patients from external datasets in healthcare centers in Argentina and France. The performance results of WARN for the prediction of AF episodes are summarized in Table 2. Next, we analyze in more detail the performance of WARN in each of these datasets.

**Table 2:** Summary of Key Performance Metrics for WARN.

| Dataset | Accuracy (%) | AUROC | AUPRC | Predicted Time Horizon |  |
| --- | --- | --- | --- | --- | --- |
|  |  |  |  | Mean | Median |
| Test (RRI) | 82.7 | 0.90 | 0.88 | 30.8 min | 38.0 min |
| Test (ECG) | 82.4 | 0.95 | 0.96 | 32.5 min | 43.4 min |
| External Centers | 75.0 | 0.80 | 0.73 | 31.8 min | 41.3 min |

### RRI data

Figure 2 presents the results for the RRI test dataset. The threshold of 0.57 was selected from the validation data as the optimal value that balances the tradeoff between accuracy and predicted time horizon (see Supplementary Material, Section S6). Figure 2 also includes results for two other thresholds to contrast tradeoffs between sensitivity and specificity. Fig. 2b shows that, for the threshold of 0.57, AF onset is predicted at least 30min in advance for around 60% of all patients in the test cohort, while attaining relatively high performance metrics (Fig. 2c). Overall, the performance of WARN is balanced between the “danger” and “normal” classes, attaining high AUROC and AUPRC scores (Fig. 2d). Smaller thresholds tend to increase the average predicted time horizon for an AF onset across patients at the expense of smaller accuracy and a larger number of false positives (Fig. 2e). For patients at risk, the threshold could be smaller and more sensitive to reduce false negatives (Fig. 2f). Finally, WARN seems to achieve similar performance for patients in different age groups; Fig. 2a shows that AF in younger patients (less than 65 years old) can be predicted around 3min earlier than for older patients.

**Figure 2:**
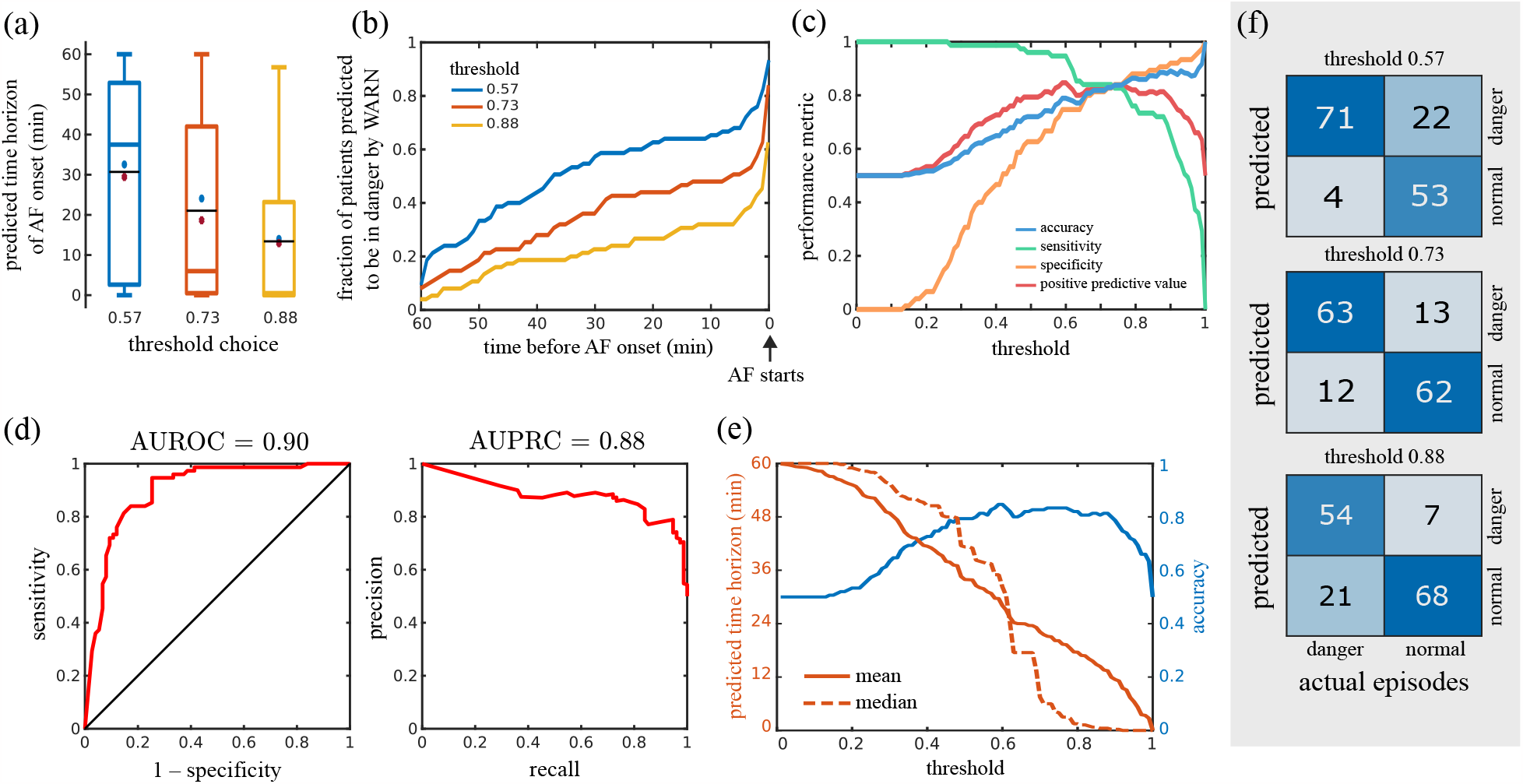
Performance of WARN on the RRI test dataset. We choose three probability thresholds (0.57, 0.73, and 0.88) to contrast tradeoffs between sensitivity and specificity. **(a)** Boxplots of the predicted time horizon (first early warning until AF onset) for different probability thresholds across all patients. Median and mean values are marked by colored and black lines, respectively. Blue circles and red asterisks represent the means of patients younger and older than 65 years, respectively. **(b)** Fraction of patients predicted to be in danger as a function of time before the AF onset for different thresholds. **(c)** Performance metrics as a function of the probability threshold. The curves cross at a threshold of 0.74 (with a value of 83.6%). **(d)** Receiver operator characteristic curve (left) and precision-recall curve (right). **(e)** Tradeoffs between the predicted time horizon and model accuracy as a function of the probability threshold. **(f)** Confusion matrices for different thresholds computed on 75 episodes of AF out of the 70 patients in the test set.

We analyzed all false predictions with the 0.57 threshold, negative and positive, to gain insight on the algorithm. Table S3 lists the observations on the incorrectly classified patients by WARN. Of the 4 false negatives, one patient had a sudden AF onset with a very stable SR beforehand; the other 3 patients had a combination of tachycardia, bradycardia, unstable base lines, and noisy signals before AF onset. Of the 22 false positives, 13 had premature atrial contractions (PACs), 5 had premature ventricular contractions (PVCs), 6 had unstable baselines, 4 had sinus tachycardia and one had atrial flutter. We noticed that the majority of these records (15 of 22) were very noisy, stressing out the necessity of treating patients’ skin with saline or disinfectant before wearing ECG devices to ensure the electrodes are well connected to the skin. Besides the noise influence, we speculate that some of these false positive events correspond to moments where the heart was close to switch from SR to AF and, for some reason, it did not. Due to a number of conditions (e.g., stress or stimulants), heart dynamics can be pushed towards the tipping point that leads to a dynamical transition from SR to AF. It is possible that, in some of the false positives, the heart was close to switch to AF, but it did not—especially for those patients with PACs (13 out of 22), which are well-known precursors of AF and highly related or causal to the occurrence of AF [42].

### ECG data

We investigate whether there is a substantial gain in performance when the CNN is trained on the original ECG data instead. Fig. S6 summarizes the performance of WARN on the test ECG data. Overall, with ECG data there was a slight improvement of model performance compared to using RRI data: the AUROC and AUPRC increased respectively by 5.5% and 9.1%, albeit the accuracy and the mean predicted time horizons were relatively similar (Table 2). Given the loss of information present in the conversion of ECG to RRI data, it is not surprising that the model trained on ECG data has improved performance compared to the model trained on RRI data. What is surprising is that the improvement was relatively small, considering the richer highly-sampled continuous-time nature of ECG data compared to the simpler low-sampled discrete-time RRI data. The achieved results show that prediction of AF onset can be efficiently performed using only RRI data; a major advantage of our work for real-time applications with wearable devices.

### RRI data from external centers

To further validate the performance of WARN on independent test datasets covering other demographics, we used ECG data collected from patients with AF from healthcare centers in Argentina (8 patients) and France (25 patients). The ECG data were first converted to RRI data. Then, using the same hyperparameters used in the WARN testing set, we obtained an accuracy of 75% and a mean (median) predicted time horizon until AF onset of 31.8min (41.31 min). Figure S7 summarizes the model performance for this external center dataset. The performance of WARN applied to this external dataset remains relatively high (close to the accuracy and mean predicted time obtained using the trained datasets), which demonstrates the potential of our method to generalize to “out-of-sample” data.

We also tested WARN’s performance on the open-access data (AFPDB) from Physionet [43], as summarized in Fig. S8. The results are shown for a balanced set of 20 AF patients and 20 healthy patients. Note that WARN was not designed for healthy patients in general since the training/test data consisted only of recordings collected from patients already diagnosed with AF. This may explain the slightly worst results of WARN on the Physionet data (accuracy of 0.7, AUROC = 0.76, and AUPRC = 0.79). Furthermore, the Physionet dataset consists only of ECG records of 30min, which led to shorter predicted time horizons for AF (mean of 12.9 minutes). Finally, it should be noted that previous models using the Physionet were not always reproducible [44].

## Discussion

This paper developed WARN, an automated prediction method for early warning of AF onset based on deep CNN and RRI signals. The method takes 30s RRI samples every 15s and computes the probability of danger of imminent AF onset. The key feature is the early and continuous rise of the probability of danger when approaching AF, providing an early warning when this probability crosses the specified threshold. On the test data (70 patients) and two external validation sets (33 patients), WARN predicted AF onset on average 31min and 33min in advance with an accuracy of 83% and 73%, respectively.

Table 3 compares the performance WARN to previous work on AF prediction. WARN is the first method to provide an early warning of AF far from onset. The prediction horizon for all previous studies is near or at AF onset. Table 3 also highlights that the dataset collected for our model training is the second larger dataset across all methods, which should increase model generalization to out-of-sample data (as validated in our analysis with external datasets).

**Table 3:** Performance comparison between WARN and previous work.

| Year | Study | Method | Nº Patients | Window length | Prediction Horizon | Accuracy | Sensitivity | Specificity |
| --- | --- | --- | --- | --- | --- | --- | --- | --- |
| 2012 | Mohebbi et al. [30] | RRI, SVM | NR | 30min | Onset | 96 | 96 | 93 |
| 2013 | Costin et al. [34] | ECG, QRS complexes | 75 | 5min | Onset | 90 | 89 | 89 |
| 2016 | Boon et al. [27] | RRI, SVM | 53 | 30min | Onset | 80 | 81 | 79 |
| 2018 | Li et al. [35] | ECG, Markov Chain | 5 | 2min | Onset | 82 | 86 | 80 |
| 2018 | Boon et al. [28] | RRI, SVM | 53 | 5min | Onset | 87 | 86 | 88 |
| 2018 | Ebrahimzadeh et al. [36] | ECG, Mixture of Experts | 53 | 5min | Onset | 98 | 100 | 96 |
| 2021 | Guo et al. [32] | RRI, XGBoost | 554 | 1h – 4h | Onset | 88 | 82 | 96 |
| 2021 | Tzoul et al. [45] | ECG, CNN | 8 | 5min | Onset | 89 | 88 | 89 |
| 2022 | Grégoire et al. [33] | RRI, CNN | 140 | 300 RRI (~5min) | 30 beats (~30s) | 66 | 80 | 53 |
| 2023 | WARN | RRI, CNN | 350 | 30s | 30.8min before onset | 83 | 95 | 70 |
| 2023 |  | ECG, CNN | 350 | 30s | 32.5min before onset | 82 | 95 | 69 |

Abbreviations: Not reported (NR), Electrocardiogram (ECG), RR interval (RRI), Convolutional Neural Network (CNN), Support Vector Machine (SVM).

WARN introduced two parameters that can be tuned by physicians depending on the clinical application: the probability threshold (danger indicator) and the moving average. These two parameters are roughly inverse to each other: lower (higher) moving averages required higher (lower) thresholds (Fig. S5). Our choice in this paper was based on a simple tradeoff decision to keep the accuracy, F1 score, and prediction horizon all relatively high. Different situations may require a higher weight on one of these objectives. For example, smaller thresholds yield more sensitive models with reduced false negatives, which could be used for high risk patients. On the other hand, higher thresholds lead to more specific models and reduced false positives, which may be more suitable to monitor AF patients with a lower incidence.

Compared with ECG data, results using RRI data showed a slight reduction in performance. On the test data, both exhibited similar accuracy of approximately 83%, with average prediction horizons of 32.5min for ECG and 30.8min for RRI data. This slight decrease in performance is compensated by the ease of continuously obtaining RRI data from easily accessible and cost-effective wearable devices like smartwatches, making them ideal for long-term monitoring. On a standard laptop computer, the total computational time spent on each sliding window was around 100ms. This is considerably less than 15s, which is the time until the next window, making it feasible to implement WARN in smartphones to process the streamed data from a smartwatch in real-time [46]. For instance, the deep-learning model used in this paper, EfficientNetV2, can be adapted to mobile devices through the TensorFlow Lite framework. Since smartwatches can be worn for long-term monitoring and record RRI signals, the early warning provided by WARN could potentially provide sufficient time for patients to take oral antiarrhythmic drugs on demand to prevent the onset of AF, or other targeted therapies or lifestyle interventions. Moreover, models could be retrained offline (e.g., once a day in an HPC) as new data becomes available.

### Limitations

WARN was trained on 24 hours RRI data from 280 patients. Hence, it is an “average” algorithm among those patients. With much longer time horizons on single patients, WARN could be personalized to improve its performance, and be converted into a real-time prediction algorithm that updates itself with newer incoming data. Eventually, such an algorithm could be used in a prospective study or a clinical trial to compare taking antiarrhythmic medication on demand (when patients receive warnings) versus the current approach of taking medication on a daily basis. Finally, WARN was trained on 100% Chinese patients. Although it was also tested on patients from France and Argentina with a good performance, the method can potentially be further improved if trained on specific demographics and commodities.

## Contributors

JG conceptualize the research. HZ, SD, PMB, MMR, and MC collected, curated, and annotated the data. MEG, JG, ANM, YJ, SZ, and BT developed the AI model. MEG implemented the code and validated the results. MEG, HZ, JG, ANM, JF, YY, RS, FB, CC, and XH analyzed the results. JG, YY, ANM, MMR, ZT, HD, ZT, HB, JF, XY, GW, and HTZ supervised the work. MEG, ANM, JG, HZ, JF, Y.Y., and XY wrote the original draft. All authors revised the manuscript.

## Data availability

The data was provided by the Tongji Hospital from China, the Clínica y Maternidad Suizo Argentina, and the the Groupe Hospitalier Privé Ambroise Paré - Hartmann acquired between 2014 and 2022. To protect patients privacy, the data was anonymized. The data collection teams from each center handled sample collection and anonymization. The algorithm development team received anonymized data containing only age and gender information for the subsequent algorithm development. The study design was evaluated and exempted from a full review by the Huazhong University of Science and Technology Institutional Review Board (approval number: TJ-IRB20220423), and approved by the Ethics Review Panel of the University of Luxembourg (approval number: ERP 22-057 RTMonitor). All data have been obtained according to the principles of the declaration of Helsinki. Training and validation sets from Tongji Hospital will be accessible pending approval by Xiaoyun Yang. Data use is limited to non-commercial research. Readers can request access to the training and validation sets to Xiaoyun Yang. The data used as external validation of this study are available from the open-access PAF Prediction Challenge Database https://physionet.org/content/afpdb/1.0.0/.

## Code availability

Data preprocessing and segmentation were implemented using the MATLAB software. The neural network was implemented on the Keras Framework with Tensorflow backend on Python 3.7. Codes have been deposited on GitHub at https://github.com/marino-gavidia/afirmo.git.

## Acknowledgements

The authors want to thank the support from the Luxembourg National Research Fund (grant PRIDE15/10907093/CriTiCS) and the National Natural Science Foundation of China (grants 92167201 and 82100531).

## Supplementary Material of

### S1. Data description

The original dataset from Tongji Hospital (Huazhong University of Science and Technology, Wuhan, China) consists of long-term 12-lead ECG Holter from 595 patients, where each ECG is recorded in SR at baseline and includes at least one AF episode. This study was approved by the Ethical Committee of Tongji Hospital with Institutional Review Board Approval number of TJ-IRB20220423. The beginning and end of individual AF episodes were labeled by experienced cardiologists at Tongji Hospital. The records have an average duration of 22.2 ± 2.2 hours, with a sampling frequency of 128 Hz and a resolution of 12 bits. We excluded records that did not have both SR and AF episodes. The records starting from AF were also excluded since the section of ECG preceding AF cannot be segmented. We considered only AF episodes with a duration of 10min or longer. Finally, records that have significant noise artifacts before AF onset were excluded (by checking if the percentage of missing R peaks within a 5min sliding window is above a threshold of 15%). After the exclusion criteria are applied, the remaining 350 records were used in this study. The cohort was divided into two groups following a chronological order between 2014 and 2019: the first 80% (280 patients) were used for the training/cross-validation of the model (252 for training and 28 for validation) and the last 20% (70 patients) for testing (Table 1).

To externally validate the performance of WARN on independent test datasets, we considered additional ECG data collected from patients with AF from three healthcare centers in different countries: the Clínica y Maternidad Suizo Argentina (53 patients of 24h ECG), the Groupe Hospitalier Privé Ambroise Paré - Hartmann (250 patients of 24h ECG) in France, and the open-access data Atrial Fibrillation Prediction Database (AFPDB) from Physionet [1] (75 patients of 30min ECG). Applying the same exclusion criteria described above resulted in a total of 73 patients for external validation: 8 from Argentina, 25 from France, and 40 from Physionet. The Physionet database consisted of 50 healthy controls (SR) and 25 AF patients (with 30min ECG just before AF onset and 5min right after). In the Physionet database, 5 AF patients presented AF with duration shorter than 1min and 2 records from healthy controls presented AF and heavy distortion due to artifact noise, which were excluded. Overall, there were 20 records to predict AF plus an equal number of 20 randomly selected records from healthy subjects for control.

### S2. Method overview

WARN is a method for early warning of the onset of AF episodes. Figure S1 illustrates the method pipeline, which is subdivided in four stages. We provide an overview of WARN as follows.

1. The ECG Holter recordings are segmented into three classes: SR, pre-AF (the instances just before AF onset), and AF segments. The AF segments were labeled by cardiologists. The pre-AF segments are labeled as the ECG data preceding the AF onset, which are characterized by high RRI variability. The SR segments comprise the remaining data, which typically have lower variability [2]. The segmentation of pre-AF intervals is described in Section S3.
2. A sliding window in the ECG data extracts segments of 30s which are converted to RRI data. The baseline wander and interference noise from the ECG are reduced using a band-pass filter with cut-off frequencies of 0.5 and 40 Hz. For each 30s window of ECG data, we detect the R peaks using the Pan-Tompkins algorithm, which has an average error rate of about 1% [3, 4]. The interval between heartbeats (i.e., from R to R peaks) is calculated to generate the RRI signal.
3. Each 30s window of RRI data is converted to a recurrence plot. Recurrence plots are 2-dimensional representations of the recurrent states of a time-series signal, which can be used to assess the periodicity of a signal and detect dynamical transitions (such as SR to AF) in a system [5–8]. The recurrence plots generate 2-dimensional images of size 224 × 224 pixels that are appropriate for CNN models used in image classification. The generation and interpretation of recurrence plots is described in Section S4.
4. A CNN model is trained to classify the inputs (recurrence plots) into the three classes segmented above: AF, pre-AF, and SR. We implemented the EfficientNetV2, a deep CNN with 479 layers, developed by Google in 2021 [9]. The EfficientNetV2 is a modified and optimized version of EfficientNet [10], winner of the ImageNet 2019 competition [11]. The input of EfficientNetV2 are images of size 224 × 224 pixels. The last fully connected layer was modified to perform the classification among the three classes. Hence, three probabilities are output by the network: *P* (SR), *P* (pre-AF) and *P* (AF), which correspond to the probability of the input data belonging to each of the three regimes (satisfying *P* (SR) + *P* (pre-AF) + *P* (AF) = 1). The training of the deep CNN model is described in Section S5. Other machine-learning and deep-learning models were also trained and tested during the development of this project, but worst performance was obtained than the current pipeline (Section S5).

**Figure S1:**
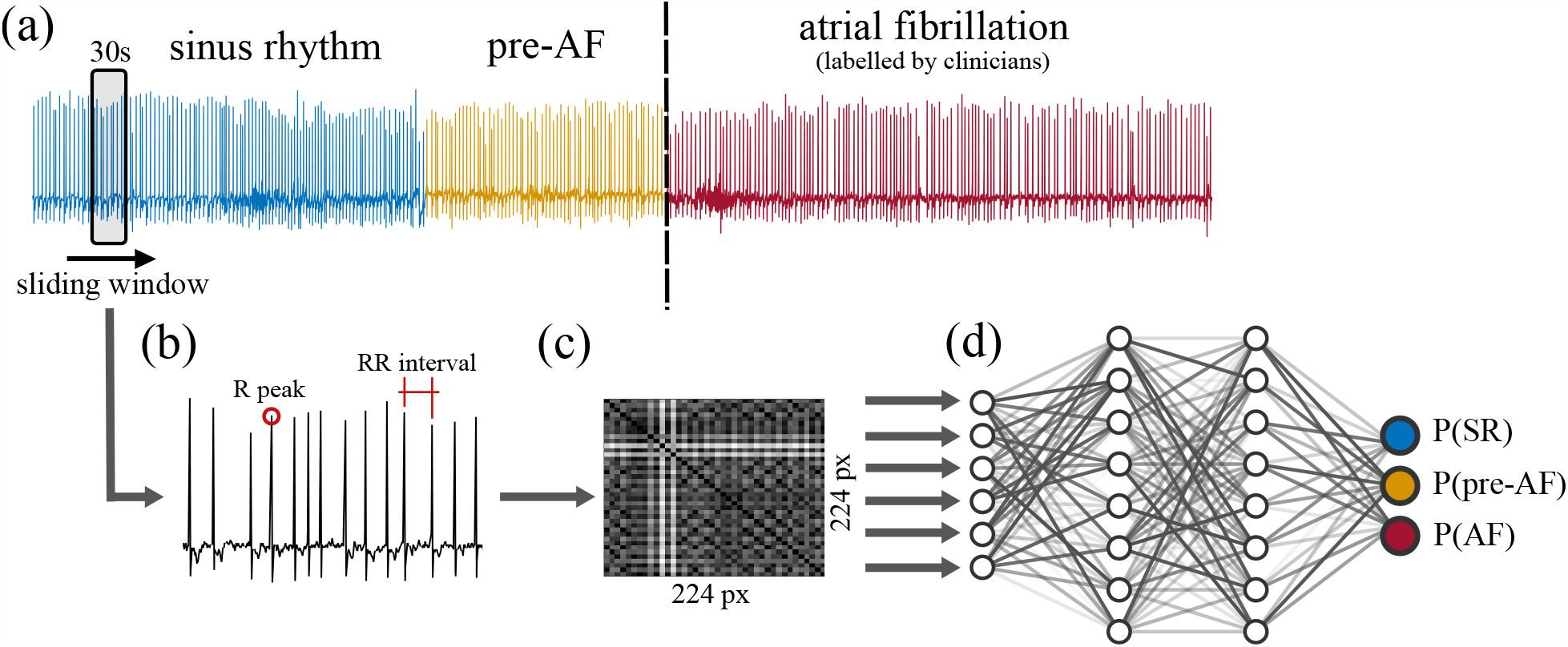
Pipeline of the first stage of WARN. **(a)** Each ECG record is split into three classes: SR, pre-AF, and AF. **(b)** The R peaks are automatically detected in a sliding window of ECG data, allowing the conversion of the ECG data to RRI data. **(c)** A recurrence plot is constructed using the RRI data window. **(d)** A deep CNN is trained using the recurrence plots as inputs. The model outputs are the probabilities that the sampled data belong to each of the three classes (SR, pre-AF, and AF).

To provide an early warning of AF, WARN computes the probability of a patient switching to AF from the outputs of the trained CNN. We define the probability of danger as *P* (danger) = *P* (pre-AF) + *P* (AF), which represents the probability that a sliding window is either in the pre-AF or AF class. Given an entire recording of RRI data (possibly converted from ECG data), our method sequentially samples a sliding window of 30s every 15s. For each new window, the recorded 30s data are converted into a recurrence plot and fed to the CNN for classification. Figure 1c illustrates the probability of danger computed by WARN for a representative example. Since the probability of danger has very high variability, we implemented a non-anticipative moving average window to filter this high-frequency noise and smooth the output (Fig. 1d). A binary early-warning indicator (“danger” or “normal”) also requires the selection of a particular threshold of the probability. These two hyperparameters, the moving average window length and probability threshold, can be optimized to maximize different performance metrics, depending on the needs of particular patients. The hyperparameter optimization is described in Section S6. The resulting hyperparameters are a moving average window size of 7 samples (corresponding to a 1.5min window) and a threshold of 0.57. This produced an accuracy of 86.7%, F1 score of 87.5%, sensitivity of 93.3%, and specificity of 80%. The mean (median) predicted time horizon until onset of AF is 31.4min (36.3min).

### S3. Pre-AF ECG segmentation

The labeling of the pre-AF ECG segments consists of five steps, illustrated in Fig. S2 for a representative patient. First, starting from the AF onset (labeled by clinicians), we select a sliding window to extract ECG samples of 5min with 30s overlapping; the sliding window moves backward in time. Second, within each 5min window, we generate a second sliding window to extract smaller samples of 30s every 5s. Third, we use the Pan–Tompkins algorithm [3, 4] to detect R waves from lead II for each 30s window and calculate the RRI. Fourth, we compute the coefficient of variation of the RRI for each 30s window and generate the corresponding histogram for each 5min window. Fifth, we analyze the evolution of the distribution of frequencies until their median is less than 0.7. The threshold of 0.7 is selected as the interception point between the distributions of frequencies of the coefficient of variation of the AF and SR regimes for all patients from the training set (Fig. S4a). Below this threshold, the heart dynamics have low variability and can be associated with SR [12]. At this fifth and last stage, the pre-AF section is segmented, including the beginning of this last window until the onset of AF. The pre-AF segments vary in length from patient to patient [12] and may also vary within multiple onsets of AF for the same patient due to morphological and electrical changes in the heart over time [13].

### S4. Recurrence plot generation

For a time-series window with *N* points, let the multi-dimensional data point *x*(*k*) *∈* ℝ^*n*^ represent the “state” of the system at time *k* = 1, …, *N*. The recurrence plot is a *N* × *N* matrix defined by the pairwise distance of all states along the recorded time-series window: *R*_*ij*_ = ∥*x*(*i*) − *x*(*j*)∥, where ∥ *·* ∥ is the Euclidean norm [14]. For a given (*i, j*)-th cell in the recurrence plot, the darker the plot (i.e., smaller *R*_*ij*_), the closer (recurrent) two states *x*(*i*) and *x*(*j*) are in the state space (see Fig. S3 for an example). For instance, darker parallel diagonal lines indicate periodicity in the state trajectory of a system, as *R*_*ij*_ decreases the closer two states are.

**Figure S2:**
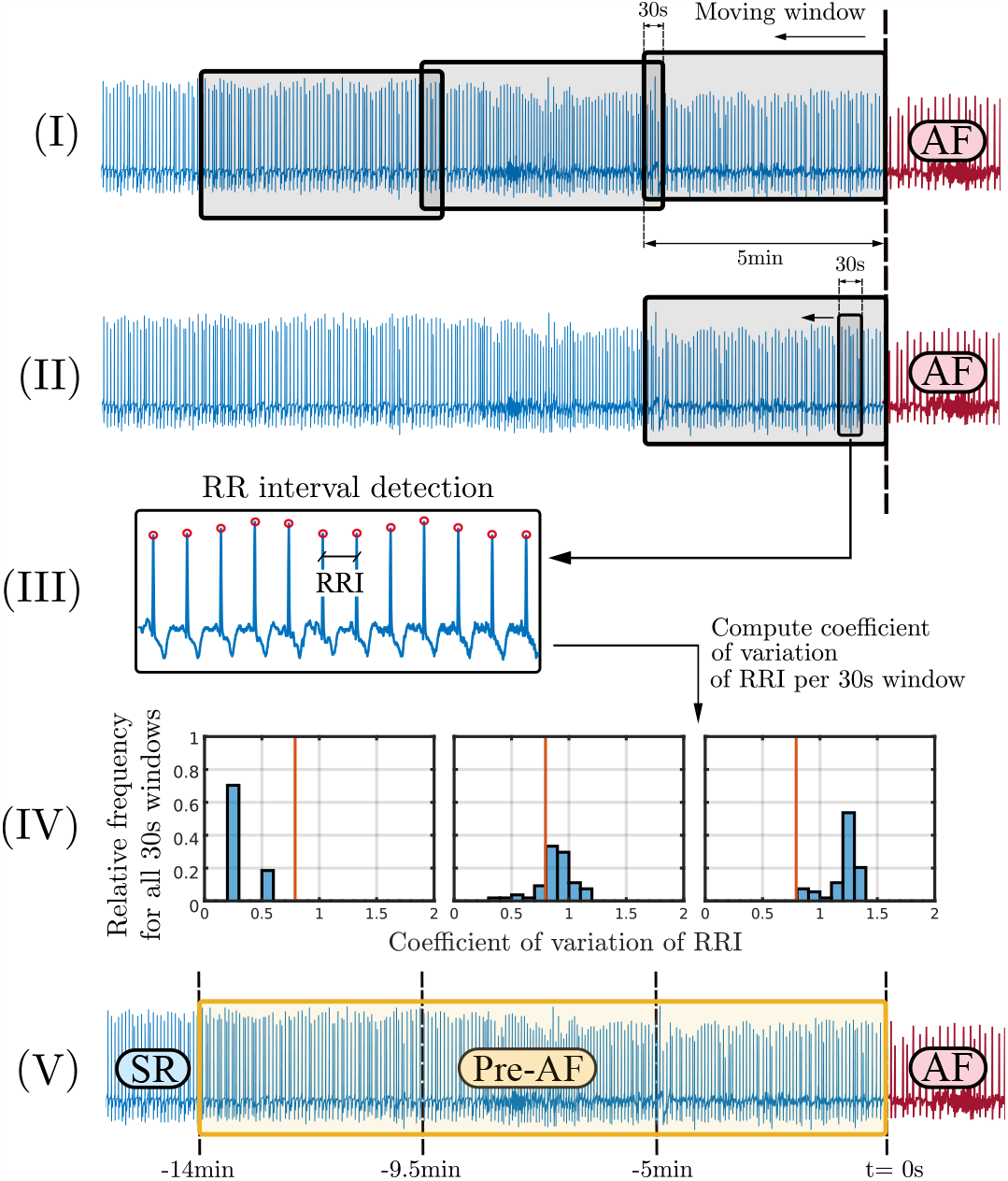
Pre-AF labeling process for a representative patient. **(I)** Starting from the AF onset and travelling back in time, a sliding window is generated to extract ECG samples of 5min with 30s overlapping. **(II)** For each 5min window, a second sliding window is generated, to extract samples of 30s every 5s. **(III)** R waves are detected for each 30 s window and the RR intervals are calculated. **(IV)** The coefficient of variation of the RR intervals of all 30s windows within a 5min window is calculated. The process is then repeated for each 5min window to generate a histogram of coefficient of variation of *RRI*. **(V)** When the histogram of *σ*_RRI_ has a median less than 0.7, the Pre-AF section is segmented from the beginning of this last window until the onset of AF. In this example, Pre-AF lasts 14min before the onset of AF.

To generate the recurrence plots in this paper, for each time window of 30s, an RRI signal is computed from the recorded ECG time-series data. Let *y*(*k*) ∈ ℝ^1^, for *k* = 1, …, *N*, be the *k*-th data point of the RRI signal, where *N* is the number of data points. The recorded time series *y*(*k*) is a 1-dimensional measurement of a (very likely) high-dimensional system. Following Taken’s theorem, we can reconstruct an attractor that preserves the structure of the original (non-measurable) state space by employing a time-delay embedding of the recorded time series [15, 16]. The embedded state vector is thus defined by *y*_*e*_(*k*) = [*y*(*k*) *y*(*k* + *τ*) *y*(*k* + 2*τ*) … *y*(*k* + (*m* − 1)*τ*)]^T^, where *m* is the embedding dimension and *τ* is the parameter of delay in data points. For the RRI data the embedding parameters are *m* = 2 and *τ* = 3, while for the ECG data we have *m* = 5 and *τ* = 6. The selected parameters *m* and *τ* correspond, respectively, to the first local minimum of the mutual information function [17] and the smallest value such that the percentage of false nearest neighbors is below 10% [18]. Finally, the recurrence plot is computed as *R*_*ij*_ = ∥*y*_*e*_(*i*) − *y*_*e*_(*j*)∥. The reader is referred to [5] for more details on recurrence plots and the choice of parameters.

Figure S3 shows recurrence plots corresponding to different regimes (SR, pre-AF, and AF). Comparing the SR and AF regimes, it is possible to observe how the number of states with high recurrence (small *R*_*ij*_) decreases on average (Fig. S3b), implying that the periodicity of the system weakens. The pre-AF segments highlight a dynamical transition with an abundant number of “cross-shaped” regions of low recurrence, which are a consequence of intermittency in the RRI signal, that is, the alternation between periodic (SR) and non-periodic (AF) regimes. We hypothesize that WARN is capable of detecting subtle patterns across all three classes, enabling it to appropriately trigger an early warning for the onset of AF.

### S5. Training of the deep CNN

WARN was trained and cross-validated on random samples from 280 patients. The CNN was trained using categorical cross-entropy as the loss function, using a batch size of 32 samples, ADAM as the optimizer [19], and stochastic gradient descent as the objective function optimizer [20]. To compensate for the class imbalance, the data was resampled and the loss function was weighted according to the ratio 3/1/2 for the SR, Pre-AF, and AF samples, respectively. The learning rate is initialized at 10^−4^ and reduced by half after the validation loss did not improve over 5 consecutive epochs. The training is terminated after 8 consecutive epochs with no improvement.

**Figure S3:**
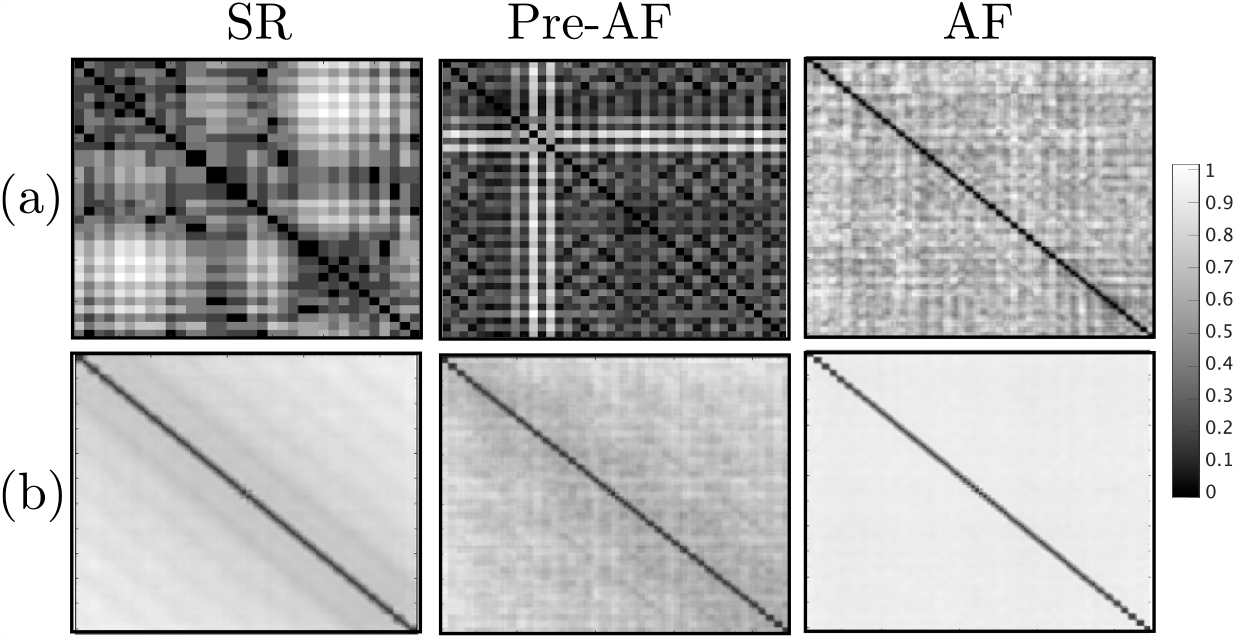
Recurrence plots, for each different segment (SR, pre-AF, and AF), generated from **(a)** a single 30 s sample and **(b)** an average of all recurrence plots generated for a representative patient.

We investigated the optimal length of the sampling window to generate the recurrence plot from the RRI data, starting from 10s up to 5min. We computed the average accuracy to predict individual samples from the 10-fold cross-validation of the EfficientNetV2 for different windows length (Table S1). The best performance was obtained using a window length of 30 seconds, as also reported in Ref. [21]. Changes in performance are associated with tradeoffs between the number of samples generated and the length of the window. The wider the window, the lower the amount of samples obtained for training, hence reducing the effectiveness of WARN to properly generalize the data. On the other hand, a smaller window length may lead to information loss [22].

**Table S1:** Optimal length of the sampling window.

| Length (seconds) | Samples ($\times 10^6$) | Accuracy |
| --- | --- | --- |
| 10 | 2.3 | 0.70 |
| 30 | 0.8 | 0.74 |
| 60 | 0.4 | 0.72 |
| 120 | 0.2 | 0.69 |
| 300 | 0.1 | 0.66 |

After fixing the sampling window length to 30s, we compared the performance of WARN with two other network benchmarks: 1-D CNN and LSTM, commonly used on arrhythmia detection and prediction [23, 24]. Using 2-D recurrence plot images as input yielded better performance than using 1-D RRI time series as input (Table S2). The proposed WARN model over-performed the benchmark networks, achieving an average validation accuracy of 0.74 and a good generalization of the data as represented by a small standard deviation of 0.03 across all 10 folds. Finally, the best model was selected for performance analysis on the test set of 70 patients. To assess performance during the second stage of WARN, the data was evenly divided between positive and negative classes, maintaining class balance and allowing fair comparisons within the confusion matrix.

**Table S2:** 10-fold cross-validation accuracy after training WARN and two benchmark networks.

| Model | 1 | 2 | 3 | 4 | 5 | 6 | 7 | 8 | 9 | 10 | Average |
| --- | --- | --- | --- | --- | --- | --- | --- | --- | --- | --- | --- |
| EfficientNetV2 | 0.78 | 0.71 | 0.73 | 0.73 | 0.73 | 0.71 | 0.75 | 0.75 | 0.76 | 0.71 | 0.74 |
| LSTM | 0.64 | 0.61 | 0.63 | 0.61 | 0.61 | 0.58 | 0.60 | 0.63 | 0.57 | 0.60 | 0.60 |
| 1D-CNN | 0.71 | 0.68 | 0.67 | 0.71 | 0.69 | 0.67 | 0.68 | 0.67 | 0.70 | 0.70 | 0.69 |

### S6. Hyperparameters optimization

To optimize the hyperparameters (moving average window length and probability threshold), the performance of WARN was evaluated on a time series of 60min of sequential data before AF onset and also far from AF (that is, in SR), selected patient-wise. We selected this time-series length since more than 70% of all pre-AF segments have a duration shorter than one hour (Fig. S4). The selection of samples far from AF is performed randomly, at least 2 hours before AF, to guarantee that the median value of the coefficients of variation of the RRI signals (computed over the selected 60min sample) is close to the median value associated with the SR distribution computed over all patients (Fig. S5a). For the validation data, it is not possible to simultaneously maximize all performance measures as expected. For example, Figs. S5a-c show that the maximum predicted time horizon until AF onset (that is, the instant of the first early warning until AF onset) is achieved at low thresholds (Fig. S5b,c). However, the accuracy is very low for those values (bottom of Figs. S5a). Likewise, when the accuracy is maximized at 88.3%, the predicted time horizon is relatively short.

To achieve a tradeoff in the validation set, we searched for hyperparameters where the accuracy, sensitivity, specificity, and F1 score are all greater than 80%, and mean and median predicted time horizon are above 30min. There was a total of 34 hyperparameters satisfying this criterion. We selected the smallest moving average window, since this leads to lower computational and memory usage in smart devices, and, among those, the one that maximizes accuracy, F1 score, and mean and median predicted time horizon. The resulting hyperparameters are a moving average window size of 7 samples and a threshold of 0.57.

For the case in which the CNN was trained on recurrence plots generated from ECG data (see section “Performance on ECG data”), we followed the same procedure to optimize hyperparameters. This led to 8 hyperparameter combinations that satisfy the criterion. The smallest moving average is 6 samples (corresponding to 1.25min windows) and the threshold is 0.48, maximizing accuracy, F1 score, and mean and median predicted time horizon. The mean predicted time horizon is 32.5min and the median 43.4min (Fig. S5d-f).

**Figure S4:**
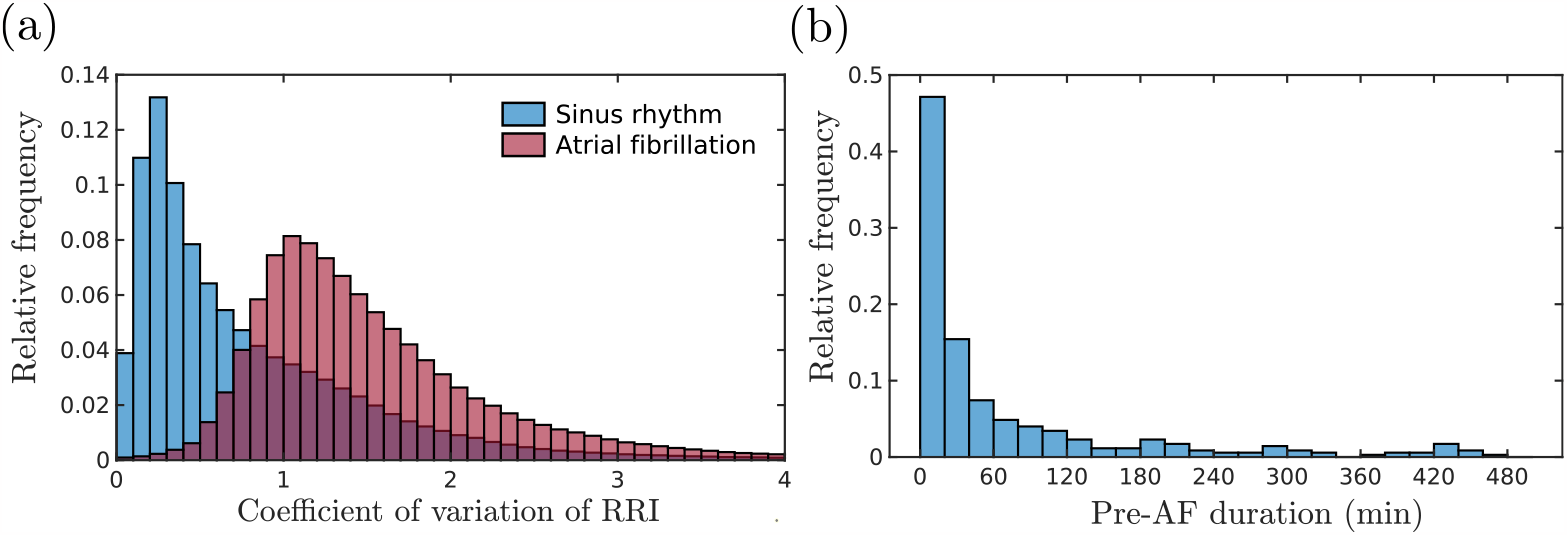
**(a)** Distributions of the coefficient of variation of the RRI for all patients from the training set, split by SR and AF regimes (as labelled by clinicians). **(b)** Distribution of Pre-AF length of for patients.

**Figure S5:**
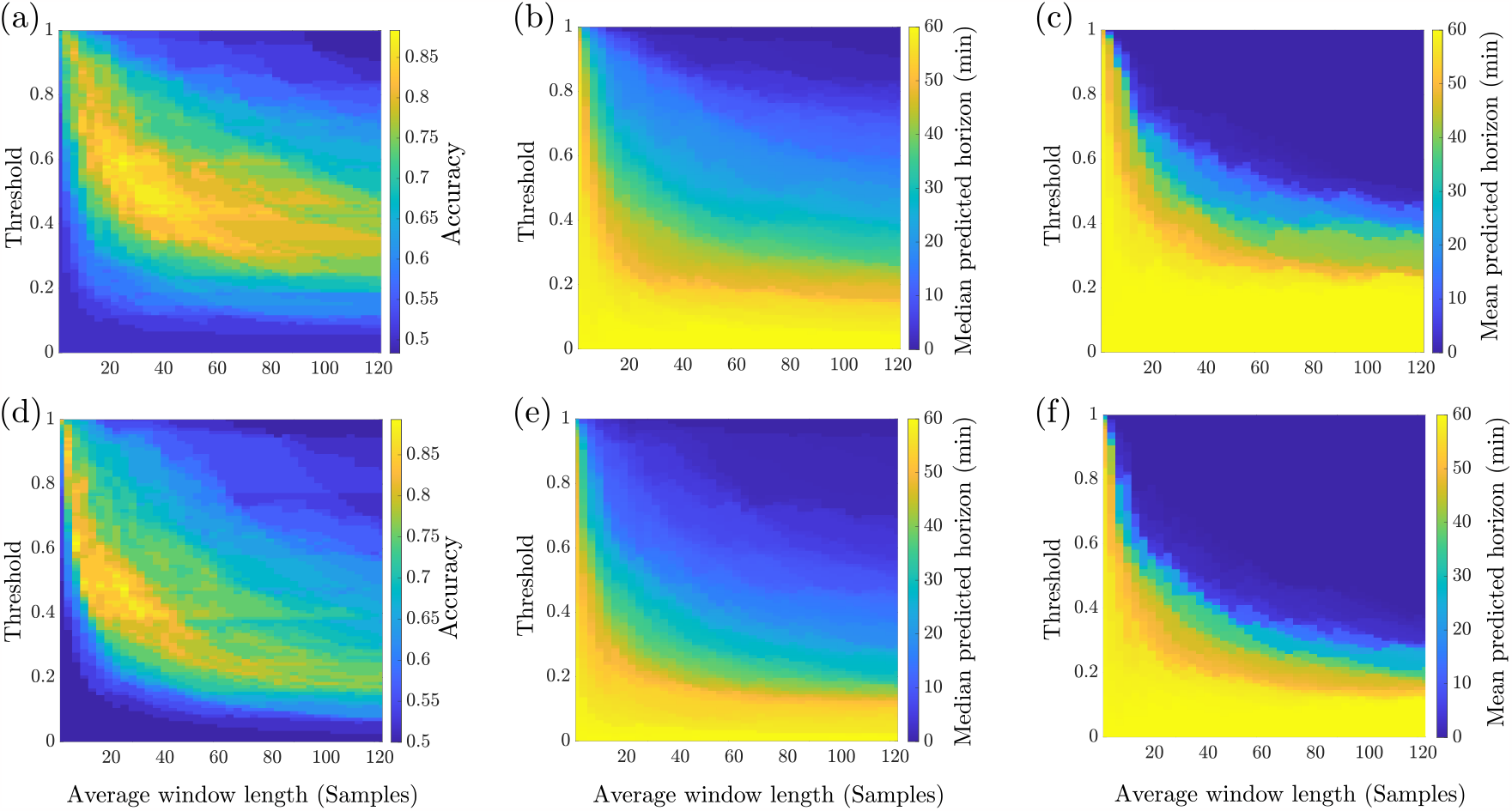
Tradeoffs on the validation data. **(a)** Model accuracy, **(b)** mean and **(c)** median of the predicted time horizon before AF onset on the validation set of the RRI data as a function of the probability threshold and the size of the moving average window that smooths the probability of danger. **(d, e, f)** Same plots computed for the ECG data.

### S7. Supporting analysis

This section contains supporting figures and table for the performance analysis of WARN.

**Table S3:** Observation on misclassification by WARN in the test dataset.

| Sample | False Negatives |
| --- | --- |
| 1 | Very stable rhythm, only SR |
| 2 | Tachycardia, bradycardia, and noise |
| 3 | Tachycardia, unstable baseline and noise |
| 4 | Unstable baseline |
| Sample | False Positives |
| 1 | Atrial flutter |
| 2, 6, 14, 19 | Noise and unstable baseline |
| 3, 8 | Multiple PVCs and PACs |
| 4, 15 | PACs and noise |
| 5 | PACs and sinus tachycardia |
| 7 | Sinus tachycardia coupled with PACs and noise |
| 9, 10, 16 | Noise |
| 11, 17, 22 | PVCs, PACs and noise |
| 12 | Sinus tachycardia, multiple PACs and noise |
| 13 | Sinus tachycardia and unstable baseline |
| 18 | Multiple PACs and noise |
| 20 | PACs, unstable baseline and long RR intervals |
| 21 | PACs |

**Figure S6:**
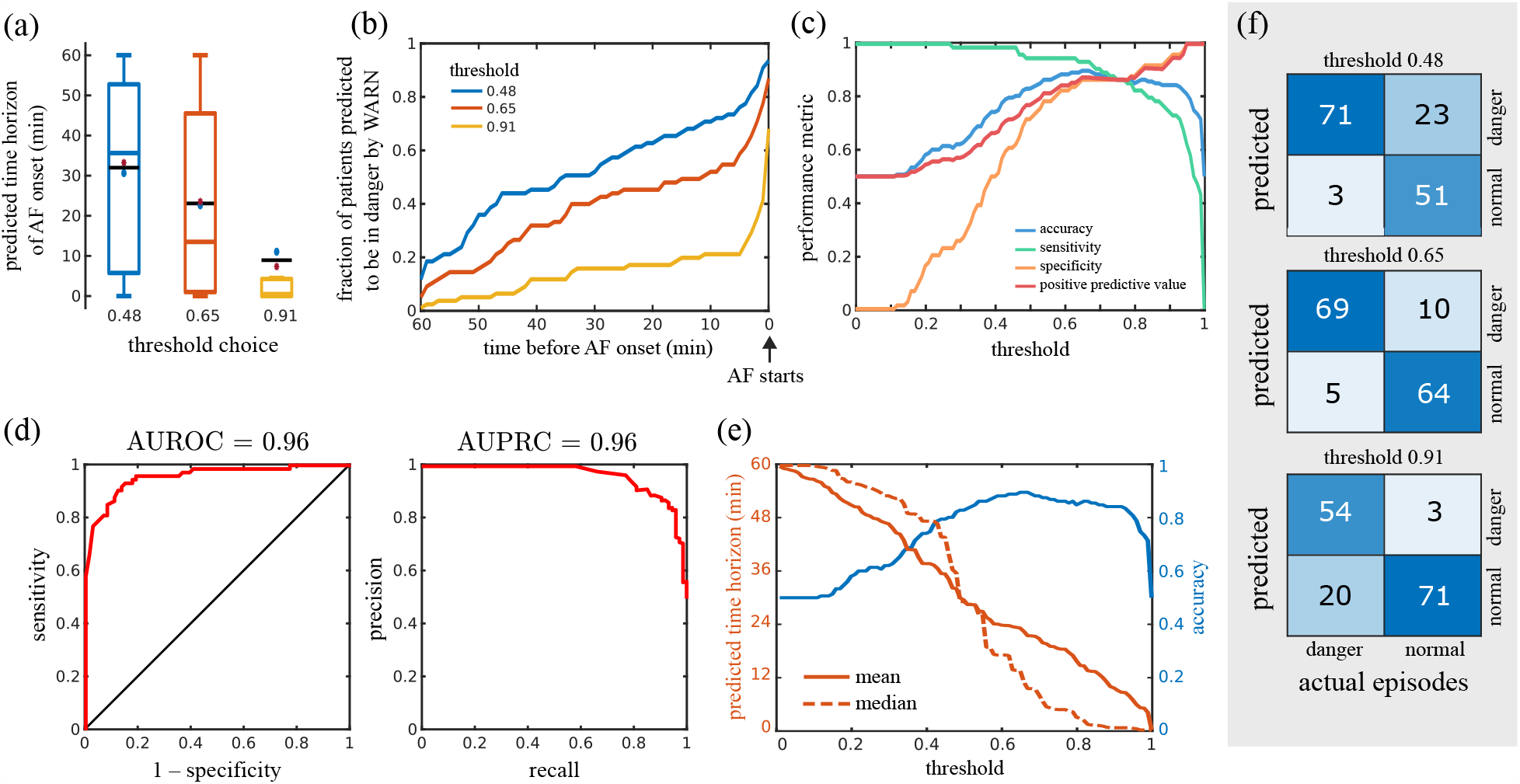
Performance of WARN on the test ECG dataset. **(a)** Boxplots of the predicted time horizon until AF onset for different probability thresholds across all patients. Median and mean values are marked by colored and black lines, respectively. Blue circles and red asterisks represent the means of patients younger and older than 65 years, respectively. **(b)** Fraction of patients predicted to be in danger as a function of time before the AF onset for different thresholds. **(c)** Performance metrics as a function of the probability threshold. **(d)** Receiver operator characteristic curve (left) and precision-recall curve (right). **(e)** Trade-off between the predicted time horizon and the model accuracy as a function of the probability threshold. **(f)** Confusion matrices for different thresholds.

**Figure S7:**
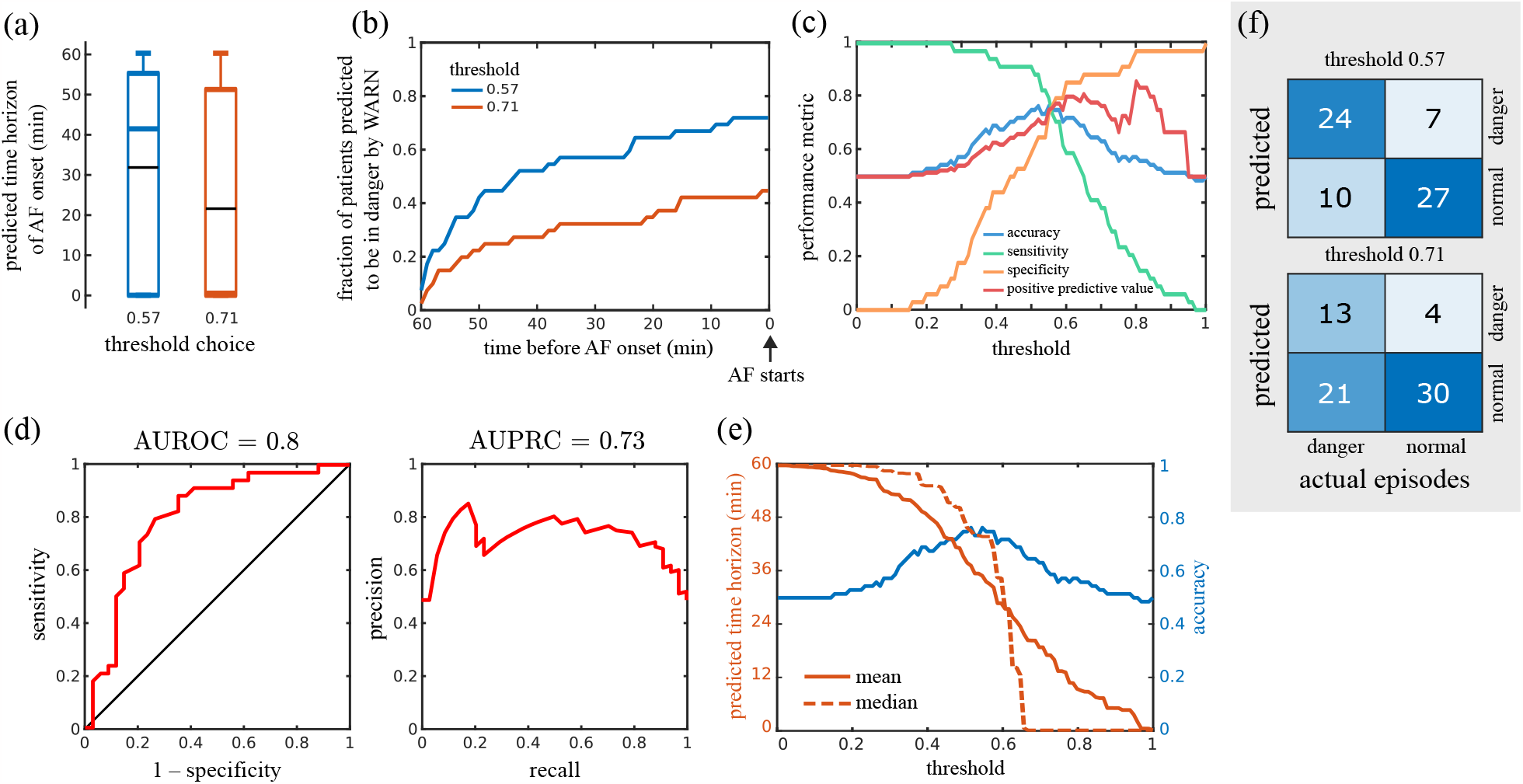
Performance of WARN on the external centers dataset. **(a)** Boxplots of the predicted time horizon until AF onset for different probability thresholds across all patients. Median and mean values are marked by colored and black lines, respectively.**(b)** Fraction of patients predicted to be in danger as a function of time before the AF onset for different thresholds. **(c)** Performance metrics as a function of the probability threshold. **(d)** Receiver operator characteristic curve (left) and precision-recall curve (right). **(e)** Trade-off between the predicted time horizon and the model accuracy as a function of the probability threshold. **(f)** Confusion matrices for different thresholds.

**Figure S8:**
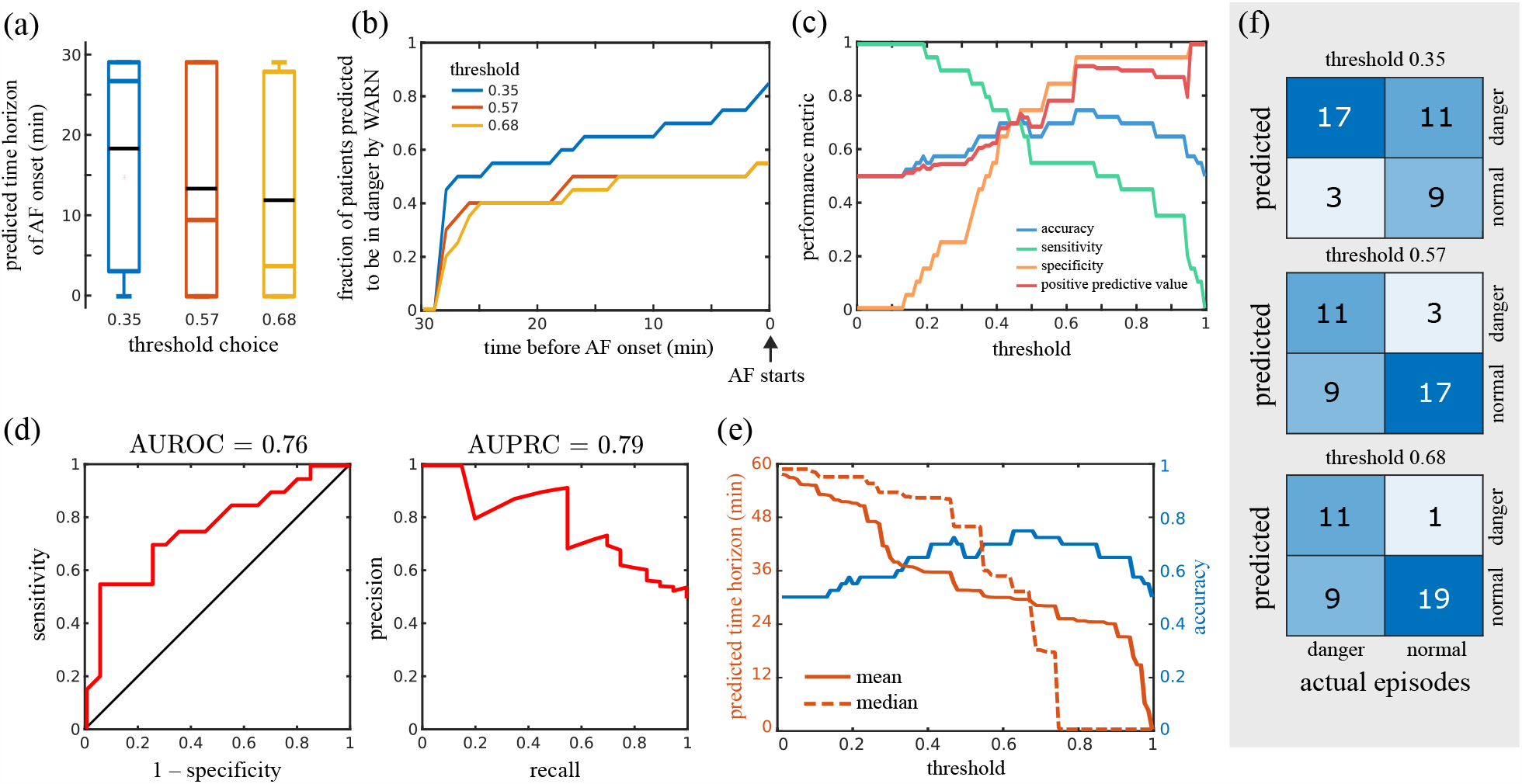
Performance of WARN on the Physionet challenge dataset. **(a)** Boxplots of the predicted time horizon until AF onset for different probability thresholds across all patients. Median and mean values are marked by colored and black lines, respectively.**(b)** Fraction of patients predicted to be in danger as a function of time before the AF onset for different thresholds. **(c)** Performance metrics as a function of the probability threshold. **(d)** Receiver operator characteristic curve (left) and precision-recall curve (right). **(e)** Trade-off between the predicted time horizon and the model accuracy as a function of the probability threshold. **(f)** Confusion matrices for different thresholds.

